# Mitral annular plane systolic excursion to left atrial volume ratio – a strainless relation with left ventricular filling pressures

**DOI:** 10.1101/2024.02.13.24302782

**Authors:** Thomas Lindow, Hande Oktay Tureli, Charlotte Eklund Gustafsson, Daniel Manna, Björn Wieslander, Per Lindqvist, Ashwin Venkateshvaran

## Abstract

**Purpose:** Left atrial reservoir strain (LASr) offers diagnostic and prognostic value in patients with heart failure. However, LASr may be technically challenging and is not available to all clinical echocardiographers. Since LASr is a consequence of left atrial (LA) stretch during apical descent of the mitral annulus, we hypothesized that a ratio between mitral annular plane systolic excursion (MAPSE) and LA volume (LAV) may offer similar diagnostic value as LASr. We aimed to investigate the relationship between MAPSE/LAV and LASr and evaluate the diagnostic performance of MAPSE/LAV to identify patients with elevated LV filling pressure.

**Methods:** MAPSE/LAV and LA strain measures were obtained in patients referred for echocardiography due to aortic stenosis, and in patients who had undergone clinically indicated right heart catheterization (RHC) with simultaneous echocardiography.

**Results:** In 93 patients with moderate aortic stenosis, MAPSE/LAV was moderately correlated with LASr (r=0.57) but was lower in patients with elevated compared to normal LV filling pressure by echocardiography (0.11 vs. 0.16 mm/mL, p<0.001). In 72 patients who had undergone RHC and simultaneous echocardiography, MAPSE/LAV and LASr correlated weakly with pulmonary artery wedge pressure (PAWP) (r=-0.44 and r=0.37). MAPSE/LAV was lower in patients with elevated (>15 mmHg) vs. normal PAWP (0.14 mm/mL vs. 0.27 mm/mL). Accuracy for detection of elevated PAWP was similar for MAPSE/LAV (area under the curve MAPSE/LAV: 0.75 [0.58–0.92] and LASr: 0.75 [0.57–0.90]).

**Conclusions:** Despite a moderate correlation with LASr, MAPSE/LAV provided similar diagnostic value as LASr regarding LV filling pressures as determined by echocardiography and RHC.

## Background

In recent years, peak left atrial longitudinal strain (LASr) has emerged as an accurate diagnostic and prognostic tool in the assessment of patients with increased left ventricular filling pressures [1–8]. However, strain measurements require advanced software and the technique is afflicted by vendor variability [9, 10]. Since longitudinal strain measurements are determined by basic volumetric changes, information obtained through strain techniques may be available from conventional measurements acquired during standard examinations.

Surrogates of strain provide undeniable advantage in resource-limited clinical environments, where technology or expertise are unavailable.

The motion of the atrioventricular plane, driven by ventricular contraction, results in a reciprocal increase of LA volume [11]. Strain-based measurements of (LA) reservoir function (LASr), i.e. the filling and stretching of the LA, is determined by the LA size and its increase during decent of the mitral annular plane in ventricular systole [12, 13], represented by mitral annular plane systolic excursion (MAPSE), Fig. 1. Thus, LASr can be described by relating the change in LA size during filling (MAPSE) to the absolute LA size. MAPSE is an easily acquired, reliable measure feasible even when image quality is suboptimal, and is an independent prognostic marker in heart failure [14, 15]. LA size is included as a standard measurement in echocardiographic examinations [16]. We hypothesized that a ratio between MAPSE and LA volume (LAV) could offer similar diagnostic value as LASr, and thus provide estimates of LA reservoir function even in the absence of speckle-tracking LA strain applications. Therefore, we aimed to describe the correlation between MAPSE/LAV and LASr, and to describe interobserver variability for both measures. Further, we aimed to determine the diagnostic value of MAPSE/LAV, in comparison to LASr, regarding detection of elevated left ventricular filling pressure.

**Figure 1.**
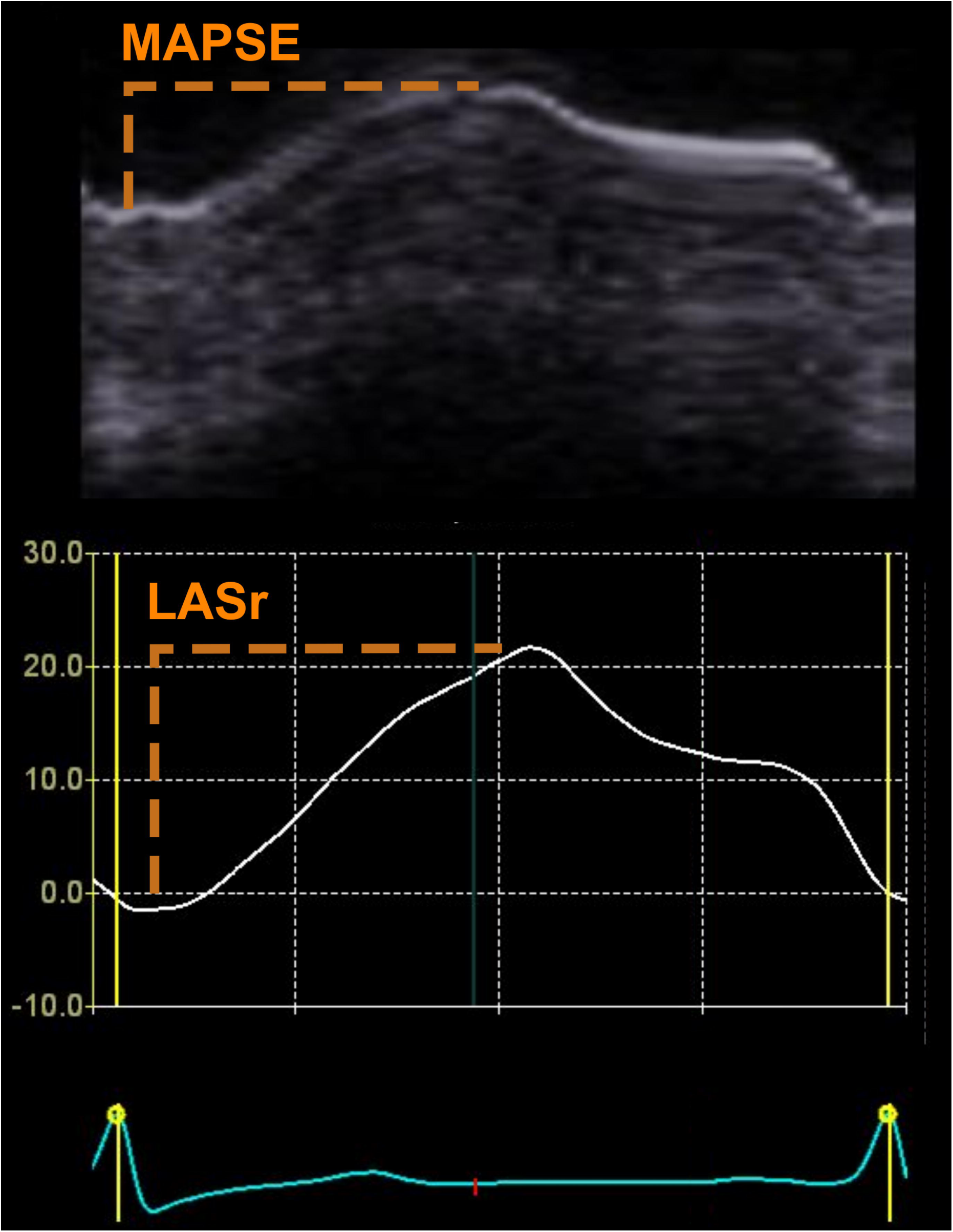
Mitral annular plane systolic excursion (MAPSE, upper panel, orange) and left atrial reservoir strain (LASr, lower panel, orange) measurements from echocardiography. Notice the close relation between MAPSE curves and LASr. The amplitude of the LASr depends on the left atrial size.

## Material and methods

Two datasets were used for this study and ethical approvals were obtained from the Human Research Ethics Committees for each cohort, respectively.

### Noninvasive evaluation of the correlation between MAPSE/LAV and LASr – Cohort 1

The correlation between MAPSE/LAV and LASr, as well as interobserver variability, were initially studied in an existing dataset including 100 patients with a clinical referral for echocardiography due to moderate aortic stenosis (aortic valve peak velocity >3.0 to <4 m/s) [17]. In the original study, these patients were either retrospectively identified (n=70) from a set starting a date and consecutively included dating backward, or prospectively (n=30) included based on the presence of a moderate aortic stenosis with the aim to evaluate interobserver variability in aortic stenosis measurements. Patients with non-sinus rhythms were excluded.

#### Echocardiography

A comprehensive transthoracic echocardiographic exam was performed in all patients using a Vivid E9 system (GE Medical Systems, Horten, Norway). Offline analyses were performed using commercially available image analysis software (EchoPAC, General Electric, Waukesha, Wisconsin, USA), and standard measurements were obtained [18]. For the purpose of this study, MAPSE and LA strain measurements were obtained by two observers. LA strain was measured on a single-beat in both 4- and 2-chamber images using the semi-automatic Automated Functional Imaging Left Atrium (AFI) tool in EchoPAC. LA contours were extrapolated across pulmonary veins and the LA appendage orifice. Zero reference strain was set at end-diastole [19]. MAPSE measurements were performed on 4-chamber images on which reconstructed M-mode from B-mode recordings of the medial and lateral mitral annular points, and measured as the vertical excursion of the mitral annulus from end-diastole to end-systole. Since mean values of all four MAPSE measurements in 4- and 2-chamber views are similar to mean values of the septal and lateral values only, MAPSE measurements from 2-chamber views were not obtained [20, 21].

LA volumes were determined through delineation of the LA in 4- and 2-chamber views at end-systole along the inner contours of the LA, excluding pulmonary veins and LA appendage, with a straight line along the mitral annulus, and calculated based on the modified Simpson’s biplane method [18]. MAPSE/LAV was then calculated as the mean of the septal and lateral MAPSE divided by the unindexed LA volume for all patients.

Patients with poor image quality, defined as at least two out six non-visualized LA segments from either the 4- or 2-chamber views or obvious foreshortening of the LA in either view, were excluded (n=7).

For the purpose of mirroring clinical reality when determining interobserver variability, no explicit measurement instructions were provided to the two observers, apart from baseline knowledge of echocardiographic guidelines for standard acquisitions, and no mutual agreement between observers were made prior to the commencement of the study. Also, both observers could freely select from the available images which images to include in their measurements. Of note, Observer 1 is an experienced reader of echocardiogram (TL; consultant, > 15 years of experience of clinical echocardiography) and Observer 2 a less experienced reader (CEG; biomedical scientist).

Echocardiographic assessment of left ventricular filling pressures was determined according to the American Society of Echocardiography(ASE)/European Association Cardiovascular Imaging (EACVI) guidelines based on the presence/absence of LA enlargement, increased tricuspid regurgitation velocity, reduced septal or lateral tissue doppler myocardial velocities (e’), and mitral peak early velocity (E)/e’ ratio [16]. In addition, quantitative estimation of left ventricular filling pressures was performed according to a recently described non-invasive method (ePAWP), which is based on LAV indexed to body surface area (LAVI), E and pulmonary vein systolic velocities (ePAWP = 0.179 × LAVI + 2.672 × mitral E/PVs + 2.7, in which ePAWP is given in mmHg, LAVI in mL/m^2^, and mitral E and PVs in the same units of velocity (e.g. both in m/s)) [22].

### Invasive assessment of the correlation between MAPSE/LAV, and LASr respectively, to PAWP – Cohort 2

To determine the correlation between MAPSE/LAV and PAWP, and LASr and PAWP, a second cohort of patients who had undergone clinically indicated right heart catheterization (RHC) and an echocardiographic examination within 1 hour (median (range) 0 (0-1) hours)) of the RHC at Umeå University Hospital, Sweden, between 2010 and 2015 (*n* = 154) [23] was included. Patients with non-sinus rhythm or at least moderate mitral regurgitation were excluded. Echocardiographic measurements were performed as described above for Cohort 1. *Right heart catherization* RHC was performed using a Swan–Ganz thermodilution catheter inserted through the right internal jugular vein, a medial cubital vein, or the right femoral vein. Pulmonary artery wedge pressure (PAWP) was defined as the mean PAWP and recorded at end-expirium during spontaneous breathing. Increased PAWP was defined as mean PAWP > 15 mmHg [24]. Main and/or contributing diagnoses for this cohort are presented in Supplements (Table S1).

#### Statistical analysis

Data are presented as mean±standard deviation (SD) or median (interquartile range) based on normal distribution. Normality was assessed using the Shapiro Wilk test. Correlations between LASr and MAPSE/LAV, LASr and PAWP, MAPSE/LAV and PAWP, respectively, were described using the Pearson correlation coefficient. A multivariable linear regression analysis was performed to describe the association between MAPSE/LAV, and LASr, respectively, with PAWP after adjusting for age, sex and LVEF. LAV was not included in the multivariable since this information is included in MAPSE/LAV. Also, GLS were not included given the strong relation to MAPSE. Differences in means between multiple groups were performed using the analysis of variance (ANOVA) test. Differences in median between groups were assessed using the Wilcoxon Rank Sum Test or the Kruskal Wallis test when comparisons of more than two groups were performed. Interobserver variability was described a the mean±SD difference, and coefficients of variation (SD of the differences between observer measurements/mean of observer measurements × 100 with 95% confidence intervals).

Diagnostic performance in detection of elevated PAWP (>15 mmHg) was evaluated using receiver operating characteristics (ROC) analysis, and presented as sensitivity, specificity, accuracy, positive likelihood ratio (LR+) and negative likelihood ratio (LR-).

Statistical significance was accepted at the level of *P* < 0.05 (two-sided). Statistical analysis was performed using R version 4.2.1 (R Core Team, Vienna, Austria).

## Results

### Cohort 1 – non-invasive assessment

After excluding 7 patients due to poor image quality, MAPSE/LAV and LASr were obtained in 93 patients with moderate aortic stenosis. Baseline characteristics are described in Table 1. MAPSE/LAV was moderately correlated with LASr (r=0.57, p<0.001), Fig. 2. MAPSE/LAV was lower for patients with increased left ventricular filling pressure, both according to the ASE/EACVI algorithm and ePAWP, Table 3 and Fig. 3.

**Figure 2.**
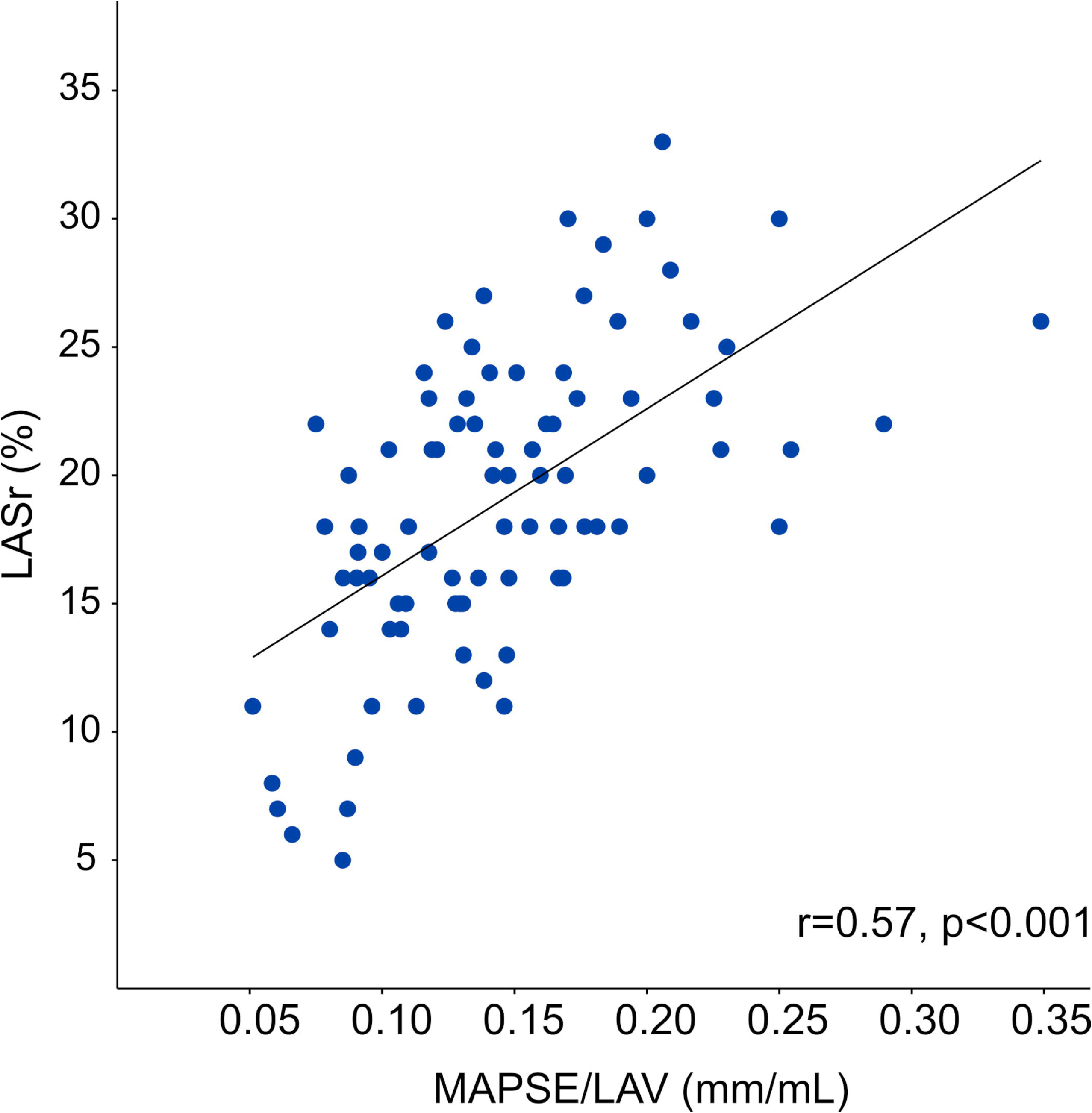
Scatterplot describing the relation between left atrial (LA) reservoir function (LASr) and mitral annular plane systolic excursion (MAPSE)/LA volume (LAV).

**Figure 3.**
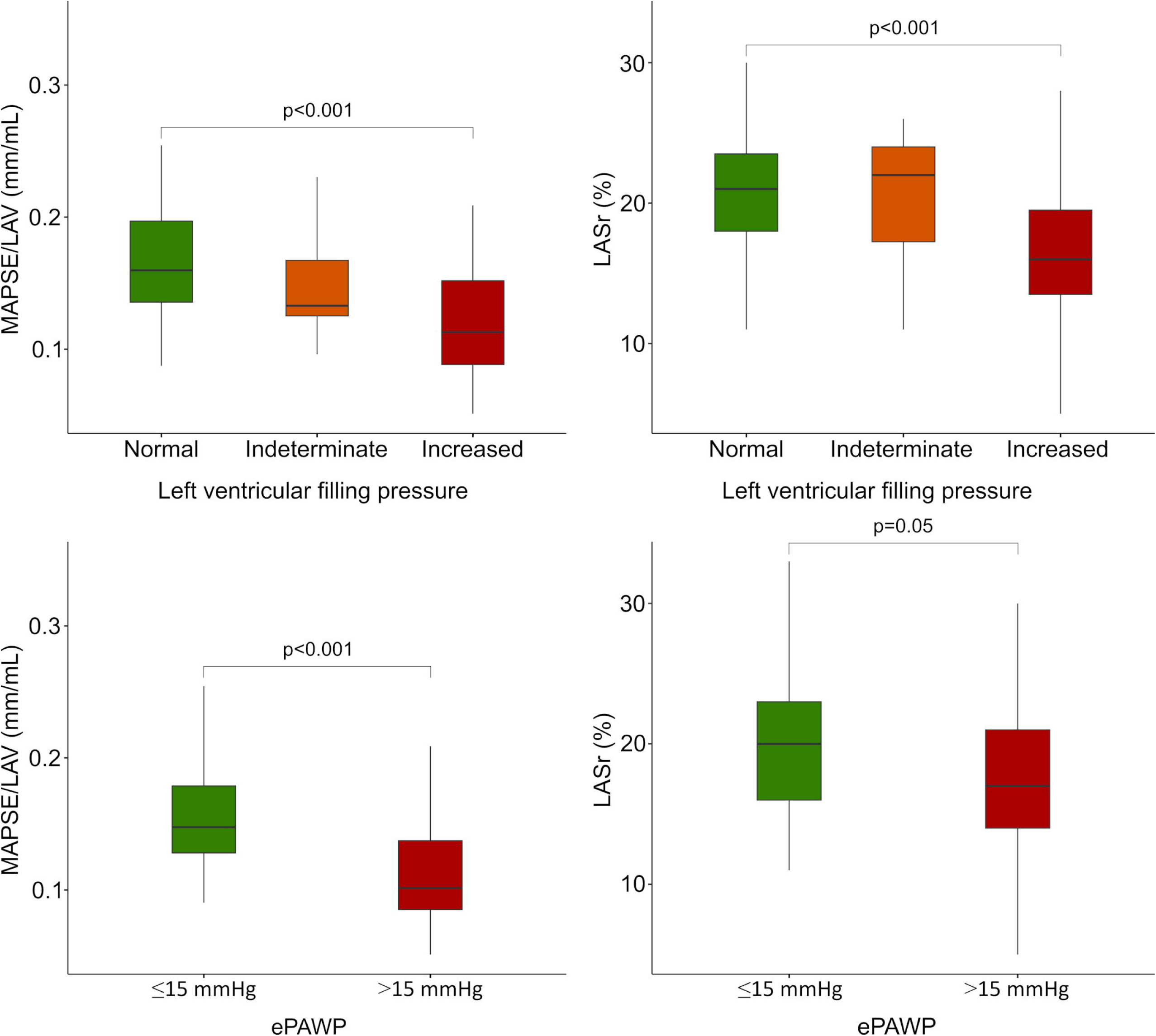
Box plots of mitral annular plane systolic excursion (MAPSE)/left atrial (LA) volume (left panels) and LA reservoir strain (LASr) (right panels) among patients with normal (green), indeterminate (orange) or increased (red) left ventricular pressure according to the ASE/EACVI guidelines for assessment of diastolic function (upper panels) and estimated pulmonary artery wedge pressure using echocardiography [22] (ePAWP; lower panels).

**Table 1.** Baseline characteristics among patients who underwent clinical echocardiography due to moderate aortic stenosis.

|  |  |  |  |  |  |
| --- | --- | --- | --- | --- | --- |
|  | Left ventricular filling pressure according to the ASE/EACVI guidelines [16] |  |  |  |  |
|  | **All** | **Normal** | **Indeterminate** | **Increased** | **p** |
| N (%) | 93 [100] | 30 [32.0] | 17 [18.3] | 41 [44.1] |  |
| Age, years | 77 [72–83] | 72[58–77] | 74 [72–83] | 83 [76–84] | 0.01 |
| Male sex, n (%) | 54 [62.8] | 24 (77.4) | 9 (56.2) | 21 (53.8) | 0.11 |
| BMI, kg/m<sup>2</sup> | 27 [26–30] | 27 [26–30] | 28 [26–32] | 28 [25–30] | 0.69 |
| BSA, m2 | 1.9±0.2 | 2.0±0.2 | 2.0±0.3 | 1.9±0.2 | 0.03 |
| *Echocardiographic measures* |  |  |  |  |  |
| LVEF, % | 60 [60–63] | 60 [60–63] | 60 [60–60] | 60 [60–65] | 0.91 |
| MAPSE, mm | 11±3 | 12±3 | 12±2 | 11±3 | 0.14 |
| LA volume, mL | 83±21 | 72±18 | 80±22 | 92±19 | <0.001 |
| LAVI, mL/m<sup>2</sup> | 43±11 | 36±8 | 41±10 | 49±10 | <0.001 |
| LV diameter, mm | 49±6 | 51±5 | 50±5 | 48±5 | 0.02 |
| LV mass, g/m<sup>2</sup> | 87±21 | 90±16 | 85±19 | 86±26 | 0.67 |
| E/A, unitless | 0.9 [0.7–1.1] | 0.9 [0.7–1.0] | 0.7 [0.6–0.9] | 1.0 [0.8–1.4] | 0.01 |
| S/D, unitless | 1.3 [1.0–1.6] | 1.3 [1.1–1.5] | 1.5 [1.2–2.0] | 1.2 [0.7–1.6] | 0.16 |
| E/e', unitless | 13 [10–17] | 10 [8–13] | 13 [9–14] | 17 [15–21] | <0.001 |
| TR velocity, m/s | 2.6 [2.5–2.8] | 2.5 [2.4–2.6] | 2.7 [2.4–2.8] | 2.8 [2.5–3.1] | 0.001 |
| AV<sub>peak</sub>, m/s | 3.4 [3.2–3.7] | 3.3 [3.2–3.5] | 3.3 [3.3–3.6] | 3.6 [3.3–3.8] | 0.02 |
| AV<sub>mean</sub>, mmHg | 28 [24–33] | 26 [23–30] | 28 [25–33] | 32 [26–35] | 0.01 |
| Data are presented as mean±standard deviation or median [interquartile range] based on normal distribution—or as n [%].<br>Abbreviations: AV<sub>peak</sub>: aortic valve peak velocity; AV<sub>mean</sub>: aortic valve mean gradient; BMI: body mass index; BSA: body surface area; LA: left atrial; LAVI: left atrial volume indexed to BSA; LVEF: left ventricular ejection fraction; LVP: left ventricular filling pressures; MAPSE: mitral annular plane systolic excursion; TR: tricuspid regurgitation |  |  |  |  |  |

**Table 2.** Mitral annular plane systolic excursion to left atrial volume ratio (MAPSE/LAV) and left atrial reservoir strain (LASr) stratified by left ventricular filling pressure according to echocardiography or right heart catherization.

|  |  |  |  |  |
| --- | --- | --- | --- | --- |
| **Table 2.** Mitral annular plane systolic excursion to left atrial volume ratio (MAPSE/LAV) and left atrial reservoir strain (LASr) stratified by left ventricular filling pressure according to echocardiography or right heart catheterization |  |  |  |  |
| *Non-invasively assessed LV filling pressure among patients with a clinically indicated echocardiography due to aortic stenosis (n=93)* |  |  |  |  |
|  | **LV filling pressure according to the ASE/EACVI algorithm [16]** |  |  |  |
|  | Normal | Indeterminate | Increased | p |
| MAPSE/LAV, mm/mL | 0.16 [0.13–0.20] | 0.13 [0.13–0.17] | 0.11 [0.09–0.15] | <0.001 |
| LASr, % | 20 [17–23] | 22 [16–24] | 16 [14–18] | 0.007 |
|  | **LV filling pressure according to ePAWP [22]** |  |  |  |
|  | Normal | Increased |  |  |
| MAPSE/LAV, mm/mL | 0.15 [0.13–0.18] | 0.10 [0.08–0.14] |  | <0.001 |
| LASr, % | 20 [16–23] | 17 [14–21] |  | 0.08 |
|  | **LV filling pressure according to invasively measured PAWP [24]** |  |  |  |
|  | Normal | Increased |  |  |
| MAPSE/LAV, mm/mL | 0.27 [0.19–0.37] | 0.14 [0.09–0.23] |  | 0.003 |
| LASr, % | 19 [12–24] | 8 [6–18] |  | 0.006 |
| Data are presented as median [interquartile range]<br>Abbreviations: LASr: left atrial reservoir strain; LV: left ventricular; MAPSE/LAV: mitral annular plane systolic excursion to left atrial volume ratio |  |  |  |  |

**Table 3.**
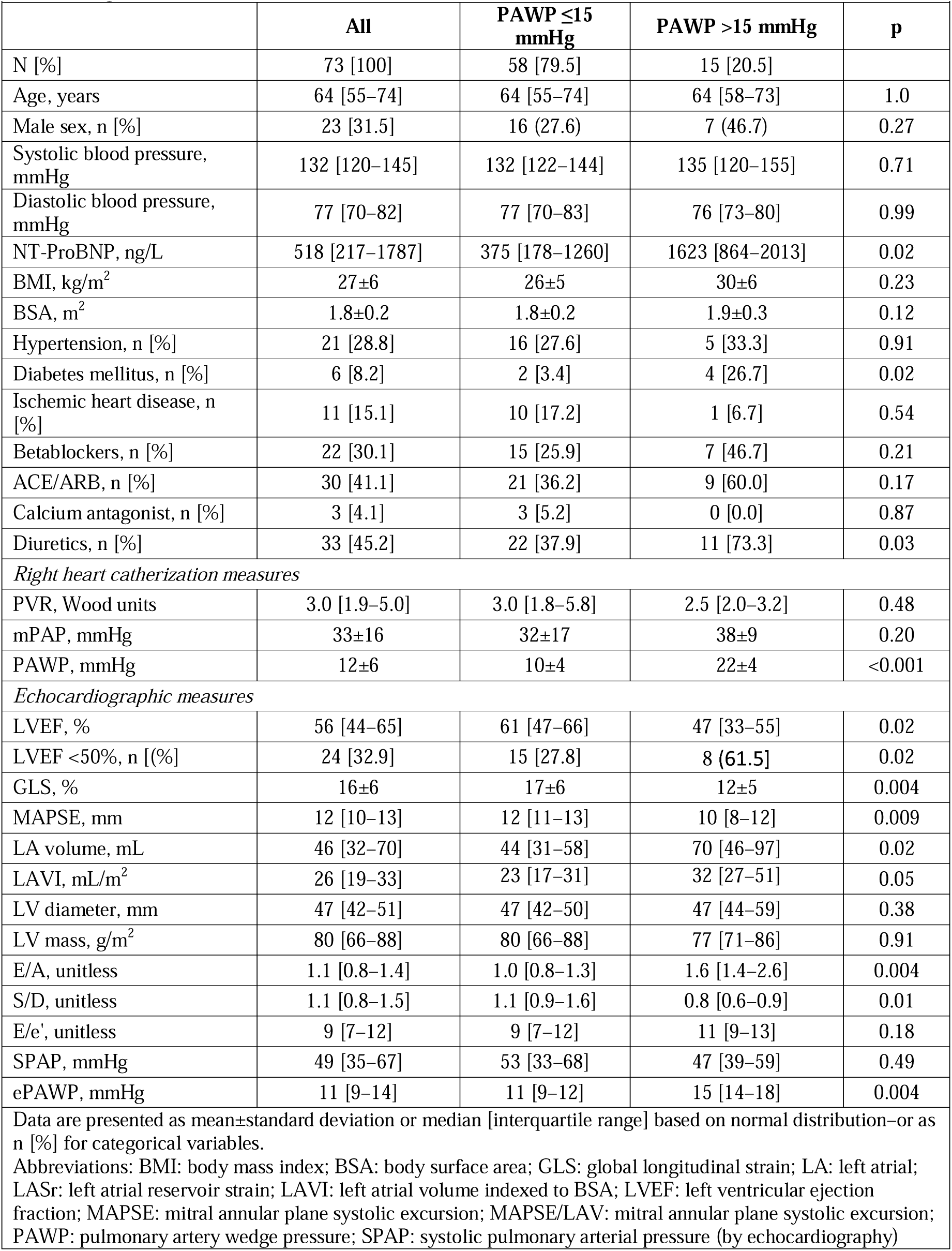
Baseline characteristics among patients who underwent echocardiography simultaneously with clinically indicated right heart catherization (n=70)

Mean difference between observers was 0.02±0.04 mm/mL for MAPSE/LAV with a coefficient of variation of 24 [19–29]%. For MAPSE and LAV alone, coefficients of variation were 14 [11–17]%, and 18 [14–22]%, respectively. For LASr mean difference between observers was 4.4±3.8%, and the coefficient of variation was 18 [14–22]%.

### Cohort 2 – invasive assessment

After excluding patients with non-sinus rhythm (n = 34), at least moderate mitral valve lesions (n = 21), missing LA strain measurements (n = 16), 72 patients who had undergone RHC and simultaneous echocardiography were finally included. Baseline characteristics are presented in Table 3. Among these, MAPSE/LAV and LASr were weakly correlated with PAWP (r=-0.44, p<0.001 and r=-0.38, p=0.001), Table 4. Similar correlations were found for MAPSE/LAV when using either the septal or the lateral MAPSE measurements only (r=0.43 for both). The association between MAPSE/LAV, and LASr, respectively, and PAWP persisted after adjusting for age, sex and LVEF (p=0.002/0.007 respectively).

**Table 4.** Correlation between MAPSE/LAV and LASr with pulmonary artery wedge pressure, left ventricular function, E/e ratio and NT-proBNP.

|  |  |  |  |  |
| --- | --- | --- | --- | --- |
|  | **MAPSE/LAV** |  | **LASr** |  |
|  | **r** | **p** | **r** | **p** |
| PAWP | -0.44 | <0.001 | -0.37 | 0.002 |
| LVEF | 0.24 | 0.045 | 0.27 | 0.02 |
| GLS | 0.19 | 0.11 | 0.49 | <0.001 |
| E/e' | -0.23 | 0.08 | -0.29 | 0.03 |
| NT-proBNP (log) | -0.33 | 0.006 | -0.29 | 0.02 |
| Abbreviations: GLS: global longitudinal strain; LA: left atrial; LVEF: left ventricular ejection fraction; MAPSE: mitral annular plane systolic excursion; NT-proBNP: N-terminal pro-B-type natriuretic peptide; PAWP: pulmonary artery wedge pressure |  |  |  |  |

MAPSE/LAV showed a stronger correlation with PAWP among patients with reduced LVEF (<50%; r=-0.54, p=0.006) than in patients with preserved LVEF (≥50%; r=-0.30, p=0.04).

Both MAPSE and LAV alone were more strongly correlated with PAWP among those with reduced LVEF (MAPSE: r=-0.49, p=0.01; LAV: r=0.55, p=0.001) compared to those with preserved LVEF (MAPSE: r=-0.17, p=0.27; LAV: r=0.29, p=0.06). Corresponding values for LASr were non-significant for both the subset of patients with reduced and preserved LVEF (LVEF <50%: r=-0.35, p=0.09; LVEF ≥50%; r=-0.26, p=0.08).

MAPSE/LAV was lower in patients with elevated (>15 mmHg) vs. normal PAWP, Table 2 and Fig. 4. Accuracy for detection of elevated PAWP was similar for MAPSE/LAV (area under the curve MAPSE/LAV: 0.75 [0.57–0.92] and LASr: 0.75 [0.57–0.90]), Fig. 5 and Table 5. The AUC for LAV, LAVI and MAPSE alone were 0.71 [0.54 – 0.88], 0.69 [0.50 – 0.88], and 0.72 [0.56 – 0.89], respectively.

**Figure 4.**
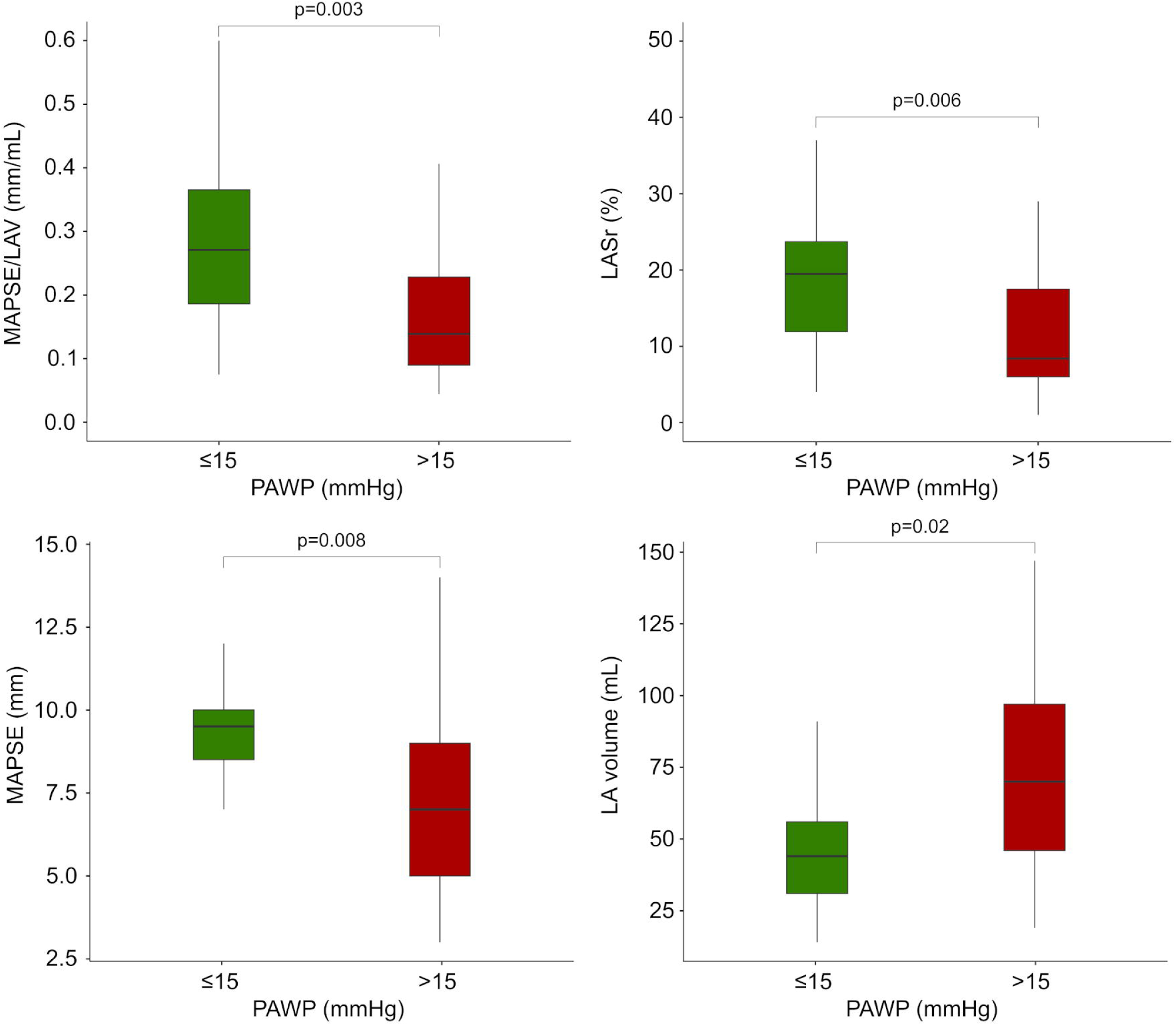
Box plots of mitral annular plane systolic excursion (MAPSE)/left atrial (LA) volume (upper left panel), LA reservoir strain (LASr) (upper right panel), MAPSE (lower left panel) and LA volume (lower right panel) among patients with normal (green) or high (red) pulmonary artery wedge pressure (PAWP) in patients who had undergone simultaneous echocardiography and right heart catheterization.

**Figure 5.**
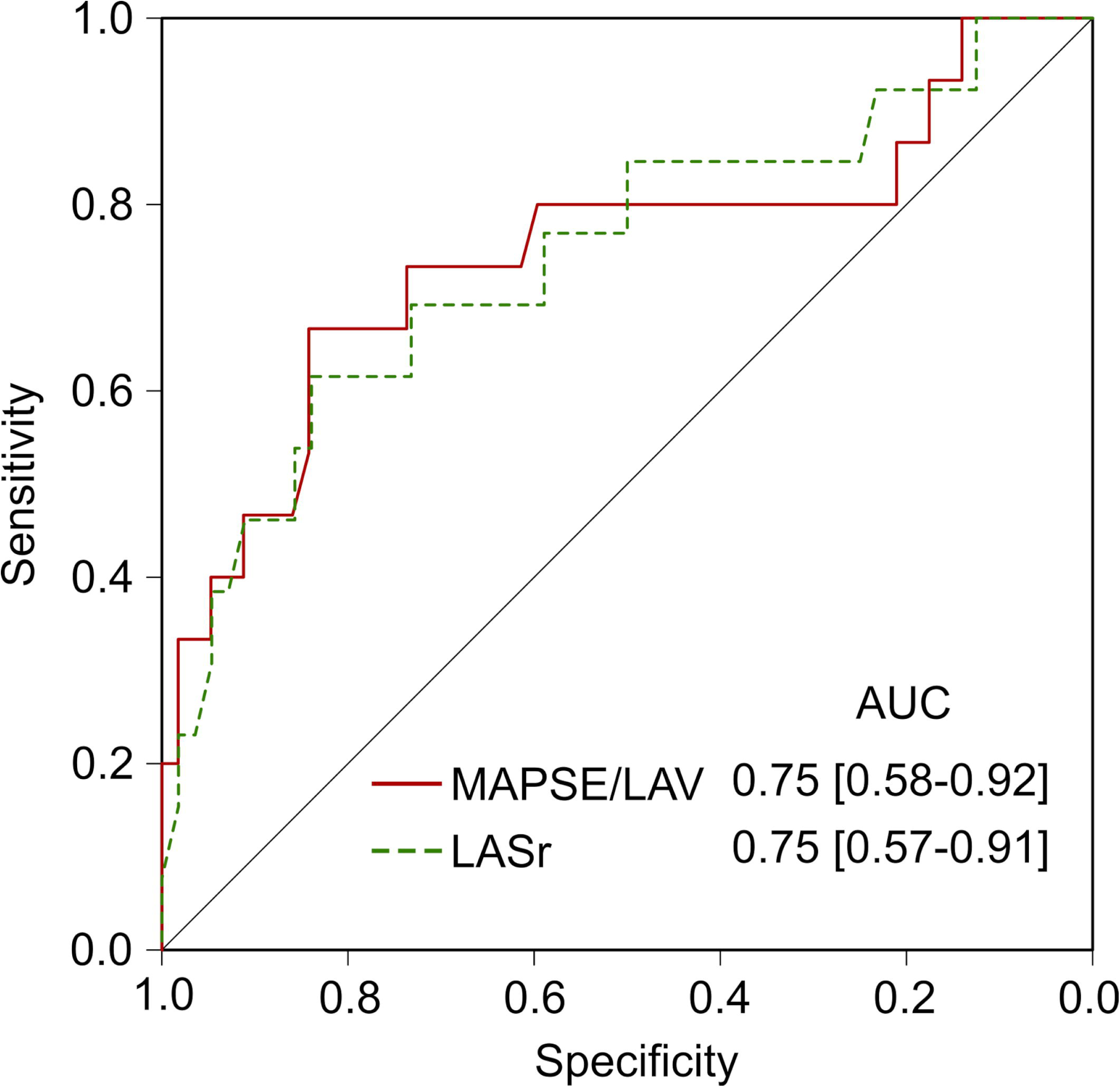
Receiver operating curve for the detection of elevated pulmonary artery wedge pressure (> 15 mmHg) according to right heart catherization.

**Table 5.** Sensitivity, specificity, accuracy, and likelihood ratios for different cutoffs for MAPSE/LAV and LASr in detection of invasively measured elevated pulmonary arterial wedge pressure (n=70).

| **MAPSE/LAV** | **Sensitivity (%)** | **Specificity (%)** | **Accuracy (%)** | **LR+** | **LR-** |
| --- | --- | --- | --- | --- | --- |
| 0.42 | 93 | 17 | 33 | 1.1 | 0.39 |
| 0.25 | 80 | 59 | 64 | 1.9 | 0.35 |
| 0.21 | 73 | 72 | 74 | 2.8 | 0.36 |
| 0.16 | 67 | 83 | 81 | 4.2 | 0.39 |
| 0.13 | 47 | 90 | 82 | 5.3 | 0.58 |
| 0.10 | 33 | 98 | 85 | 16.5 | 0.68 |
| **LASr** | **Sensitivity (%)** | **Specificity (%)** | **Accuracy (%)** | **LR+** | **LR-** |
| 24 | 92 | 23 | 36 | 1.2 | 0.35 |
| 20 | 85 | 50 | 56 | 1.7 | 0.30 |
| 18 | 77 | 59 | 62 | 1.9 | 0.39 |
| 13 | 69 | 73 | 72 | 2.6 | 0.42 |
| 9 | 54 | 86 | 80 | 3.9 | 0.53 |
| 6 | 38 | 95 | 84 | 7.6 | 0.65 |

## Discussion

A surrogate measure of LASr obtained through measurements of mitral annular displacement and LAV (MAPSE/LAV) provided similar diagnostic information as its strain-based counterpart. Since LA volumes are acquired in a standard echocardiographic examination, this measure could easily be obtained by adding MAPSE measurement to the examination. Given the prognostic value of MAPSE [14, 15, 25], in itself, the addition of this measurement thus adds both diagnostic and prognostic value to the echocardiographic examination. Both MAPSE/LAV and LASr were lower in patients with elevated left ventricular filling pressures as determined both by the ASE/EACVI algorithms, a quantitative echocardiographic PAWP estimation, and invasively measured PAWP. LASr has previously been shown to provide diagnostic information regarding left ventricular filling pressures [4, 8, 26, 27]. For example, when LASr was incorporated into the ASE/EACVI algorithm, both feasibility and accuracy in detection of elevated filling pressures were improved, and LASr could effectively be used as replacement when either E/e’ or tricuspid regurgitation velocity were unavailable [8].

Although, LAV constitutes an important part of LASr, LASr improves prediction of elevated PAWP beyond that obtained from LAV indexed-to-body surface area [28]. In this study, MAPSE/LAV and LASr showed acceptable discrimination of patients with normal or elevated PAWP (AUC ∼0.75). However, MAPSE/LAV resulted in only a small increase in diagnostic accuracy compared to its constituents alone (MAPSE and LAV, ΔAUC +0.04 and +0.03, respectively).

Importantly, both MAPSE/LAV and LASr were only moderately correlated with PAWP and neither of these measures can be used as standalone noninvasive measures of PAWP. A stronger correlation with PAWP could be observed for MAPSE/LAV in patients with reduced LVEF, in comparison to those with preserved LVEF. Although a similar pattern has been reported for LASr previously [13], this was not evident for LASr in this study. MAPSE/LAV explicitly incorporates systolic function (MAPSE), and higher PAWP could be expected with worse systolic function. However, the sample size was small and the cause for the stronger correlation for MAPSE/LAV among patients with reduced LVEF cannot be readily explored. Interobserver variability was substantial for both MAPSE/LAV and LASr, although higher for MAPSE/LAV. Possibly, this difference was caused by the semi-automatic procedure used for LA strain, in contrast to MAPSE/LAV, which likely lowers variability. However, coefficients of variation were similar for LAV and LASr, indicating differences in LA delineation between the two observers, and the addition of another measure (MAPSE) increased variability further. The high variability observed in this study contrasts previous reports of repeatability of LA strain measures, which have suggested low interobserver variability [29]. This can be explained by the use of single-beat measurements used in this study, suggesting that averaging beats over multiple cycles are needed to obtain adequate reproducibility. Also, we actively refrained from synchronizing measurement strategies between observers, to mirror clinical reality. For both MAPSE/LAV and LASr there was a systematic bias between observers, possibly caused by the different contour delineation of the LA or differences in placement of the annular detachments for both measures. The high variability of MAPSE/LAV, however, clearly limits clinical application but this finding needs further evaluation in future studies. Compared to real-time M-mode images, reconstructed M-mode images the temporal resolution is limited to that of 2D images but has been shown to provide accurate information for measurements of anatomical dimensions [30]. Still, variability could possibly be reduced by using real-time M-mode acquisitions instead of reconstructed M-mode images.

LASr is determined, to a large extent, by atrioventricular plane displacement and LA dimensions. This has been observed previously [12, 13]. Mălăescu showed excellent correlations between LV and LA strain curves across different cardiovascular pathologies. Similarly, Inoue, et al, showed LV global strain and LAV to be important determinants of LASr [13]. LV global strain describes the systolic longitudinal shortening of the LV in relation to the length of LV cavity. Thus, the main difference between MAPSE and LV global strain in relation to their association with LA strain is that LV global strain also incorporates information of LV dimensions, which is likely to be redundant. Despite this, we did not find a stronger relation between MAPSE and LASr compared to between LV global strain and LASr. Possibly because of a closer relationship of measurement techniques between the two strain modalities compared to the M-mode based measurement, or due to measurement variability.

Both MAPSE/LAV and LASr were lower in the cohort of patients with moderate aortic stenosis, compared to the cohort of patients who underwent clinical RHC. This could be explained by older patients in the aortic stenosis cohort, a higher proportion of male patients, or differences in the degree of LV disease [20, 31]. Also, reduced longitudinal function commonly occurs in patients with significant aortic stenosis [32], resulting in lower MAPSE/LAV and LASr.

### Limitations

The cohort of patients with aortic stenosis did not include individuals with suspected heart failure, representing a limitation. However, the primary objective of analysing this cohort was to describe the correlation between MAPSE/LAV and LASr, and to determine reproducibility. Nonetheless, the potential clinical utility of MAPSE/LAV requires evaluation in settings that encompass patients with suspected or confirmed heart failure.

Since zero strain is set at end-diastole, i.e. minimum LAV, a stronger correlation with LASr could be achieved by dividing MAPSE with the minimum LAV instead of the maximum LAV. Using cardiovascular imaging, minimum LAV showed a stronger correlation with LASr than maximum LAV [33]. Pragmatically, we included the maximum LAV in the MAPSE/LAV calculation since this value is included in standard echocardiographic reports and protocols. Thus, only MAPSE would be required to be added to obtain a reasonable surrogate of LASr, if needed. Future studies could be performed to demonstrate a possibly improved diagnostic accuracy of MAPSE/minimum LAV instead of the measures included here.

There is a lack of robust non-invasive reference parameters for PAWP. In the non-invasive cohort, MAPSE/LAV and LASr was therefore described both in relation to the ASE/EACVI algorithm and to ePAWP. In a previous paper, with larger sample size than the current study, ePAWP provided better diagnostic accuracy and improved prognostic value compared to the ASE/EACVI algorithm [22]. Given the limited precision of ePAWP, however, the results from this study regarding the relation between MAPSE/LAV, and LASr, and PAWP must be interpreted with caution. Nonetheless, the pattern was similar both cohorts. Although such patients were part of the invasively examined study cohort, those patients constitute a mixed population with advanced disease unlikely to be representative of the typical population in which non-invasive determination of left ventricular filling pressures is sought. Nevertheless, our findings lend support to the hypothesis that comparable diagnostic information, as derived from strain-based LA measures, can be achieved without the use of advanced software.

Patients with atrial fibrillation were excluded from this study even though atrial fibrillation is common in patients with heart failure. The main aim of this study was to determine whether simple anatomical measures could provide similar diagnostic information as strain-based techniques, and atrial fibrillation could possibly make such comparisons more difficult, since this arrythmia would introduce further variability to both echocardiographic and invasive measures [34]. The findings of this study cannot be applied to patients with atrial fibrillation. This study is further limited by small sample sizes. This is reflected, for example, in the wide confidence intervals for the AUC for both MAPSE/LAV and LASr, ranging from poor to excellent discrimination. These results, however, did not provide any evidence of improved diagnostic accuracy of LASr compared to the simple measures included in MAPSE/LAV. Inter-reader variability was assessed after re-analysis of images from the same set of images. Although both readers could freely choose which 4- and 2-chamber views to use, it should be noted that variability can increase further if new acquisitions are performed [17]. Although inter-reader variability was higher in this study than previously reported, both LASr and MAPSE/LAV should be evaluated regarding their potential in detecting change in PAWP, since such information is affected by variability in repeated measurements. Such an analysis would require sequential invasive PAWP measurements, which was not available in this study.

MAPSE is affected by regional LV dysfunction, mitral annular calcification, conduction disorders, or small areas of fibrosis [35], which is a limitation both to the results of this study and to a general use of MAPSE/LAV. However, given the importance of the longitudinal excursion of the mitral annular plane to LASr, even LASr can be affected by these conditions, and future studies of both parameters in these specific conditions may be needed. Also, including lateral, or the septal, MAPSE measurement only in the calculation of MAPSE/LAV, did not show a stronger correlation with PAWP.

LA volumes were lower in the invasive cohort compared to the non-invasively examined cohort. This can be explained by differences in patient populations, e.g. the invasive cohort consisted of patients with a clinical referral for RHC including pulmonary hypertension not due to LV disease, while the non-invasive cohort consisted of patients with moderate aortic stenosis and thus, likely, a higher degree of LV and LA disease. Although this makes generalization of absolute values for both LASr and MAPSE/LAV difficult, it does not affect the head-to-head comparison of LASr to MAPSE/LAV and their respective association with LV filling pressures.

The above-mentioned limitations indicate a need for further studies in independent cohorts before clinical implementation. The results, however, can be considered as proof-of-concept of similar diagnostic value of MAPSE/LAV and LASr, although both methods are limited as stand-alone measures of LV filling pressures.

## Conclusion

MAPSE/LAV was lower in patients with elevated LV filling pressures as determined both by echocardiography and RHC. MAPSE/LAV provided similar diagnostic information as LASr in relation to LV filling pressures, and may serve as a substitute for LASr when such measures are unavailable, but was afflicted by high interobserver variability.

## Data Availability

The data underlying this article can be shared upon reasonable request to the corresponding author.

## References

1. Lacalzada-Almeida J, Izquierdo-Gómez MM, García-Niebla J, et al. Advanced interatrial block is a surrogate for left atrial strain reduction which predicts atrial fibrillation and stroke. Ann Noninvasive Electrocardiol. 2019;24:e12632.

2. Cau R, Bassareo P, Suri JS, et al. The emerging role of atrial strain assessed by cardiac MRI in different cardiovascular settings: an up-to-date review. European radiology. 2022;32:4384–94.

3. Mandoli GE, Pastore MC, Benfari G, et al. Left atrial strain as a pre-operative prognostic marker for patients with severe mitral regurgitation. International journal of cardiology. 2021;324:139–45.

4. Cameli M, Mandoli GE, Loiacono F, et al. Left atrial strain: a new parameter for assessment of left ventricular filling pressure. Heart failure reviews. 2016;21:65–76.

5. Morris DA, Belyavskiy E, Aravind-Kumar R, et al. Potential Usefulness and Clinical Relevance of Adding Left Atrial Strain to Left Atrial Volume Index in the Detection of Left Ventricular Diastolic Dysfunction. JACC. Cardiovascular imaging. 2018;11:1405–15.

6. Park JH, Hwang IC, Park JJ, et al. Prognostic power of left atrial strain in patients with acute heart failure. European heart journal. Cardiovascular Imaging. 2021;22:210–9.

7. Rausch K, Shiino K, Putrino A, et al. Reproducibility of global left atrial strain and strain rate between novice and expert using multi-vendor analysis software. Int J Cardiovasc Imaging. 2019;35:419–26.

8. Venkateshvaran A, Tureli HO, Faxén UL, et al. Left atrial reservoir strain improves diagnostic accuracy of the 2016 ASE/EACVI diastolic algorithm in patients with preserved left ventricular ejection fraction: insights from the KARUM haemodynamic database. European heart journal. Cardiovascular Imaging. 2022;23:1157–68.

9. Mouselimis D, Tsarouchas AS, Pagourelias ED, et al. Left atrial strain, intervendor variability, and atrial fibrillation recurrence after catheter ablation: A systematic review and meta-analysis. Hellenic journal of cardiology: HJC = Hellenike kardiologike epitheorese. 2020;61:154–64.

10. Wang Y, Li Z, Fei H, et al. Left atrial strain reproducibility using vendor-dependent and vendor-independent software. Cardiovascular ultrasound. 2019;17:9.

11. Steding-Ehrenborg K, Carlsson M, Stephensen S, et al. Atrial aspiration from pulmonary and caval veins is caused by ventricular contraction and secures 70% of the total stroke volume independent of resting heart rate and heart size. Clinical physiology and functional imaging. 2013;33:233–40.

12. Mălăescu GG, Mirea O, Capotă R, et al. Left Atrial Strain Determinants During the Cardiac Phases. JACC. Cardiovascular imaging. 2022;15:381–91.

13. Inoue K, Khan FH, Remme EW, et al. Determinants of left atrial reservoir and pump strain and use of atrial strain for evaluation of left ventricular filling pressure. European heart journal. Cardiovascular Imaging. 2021;23:61–70.

14. Xue H, Artico J, Davies RH, et al. Automated In-Line Artificial Intelligence Measured Global Longitudinal Shortening and Mitral Annular Plane Systolic Excursion: Reproducibility and Prognostic Significance. Journal of the American Heart Association. 2022;11:e023849.

15. Berg J, Åkesson J, Jablonowski R, et al. Ventricular longitudinal function by cardiovascular magnetic resonance predicts cardiovascular morbidity in HFrEF patients. ESC heart failure. 2022;9:2313–24.

16. Nagueh SF, Smiseth OA, Appleton CP, et al. Recommendations for the Evaluation of Left Ventricular Diastolic Function by Echocardiography: An Update from the American Society of Echocardiography and the European Association of Cardiovascular Imaging. European heart journal. Cardiovascular Imaging. 2016;17:1321–60.

17. Manna D, Eliasson M, Bech-Hanssen O, et al. Reproducibility of Echocardiographic Measures of Aortic Stenosis Severity and Its Impact on Grading of Severity. Journal of the American Society of Echocardiography: official publication of the American Society of Echocardiography. 2023.

18. Mitchell C, Rahko PS, Blauwet LA, et al. Guidelines for Performing a Comprehensive Transthoracic Echocardiographic Examination in Adults: Recommendations from the American Society of Echocardiography. Journal of the American Society of Echocardiography: official publication of the American Society of Echocardiography. 2019;32:1–64.

19. Badano LP, Kolias TJ, Muraru D, et al. Standardization of left atrial, right ventricular, and right atrial deformation imaging using two-dimensional speckle tracking echocardiography: a consensus document of the EACVI/ASE/Industry Task Force to standardize deformation imaging. European heart journal. Cardiovascular Imaging. 2018;19:591–600.

20. Støylen A, Mølmen HE, Dalen H. Relation between Mitral Annular Plane Systolic Excursion and Global longitudinal strain in normal subjects: The HUNT study. Echocardiogr. 2018;35:603–10.

21. Støylen A, Mølmen HE, Dalen H. Regional motion of the AV-plane is related to the cardiac anatomy and deformation of the AV-plane. Data from the HUNT study. Clinical physiology and functional imaging. 2023;43:453–62.

22. Lindow T, Manouras A, Lindqvist P, et al. Echocardiographic estimation of pulmonary artery wedge pressure - invasive derivation, validation, and prognostic association beyond diastolic dysfunction grading. European heart journal. Cardiovascular Imaging. 2024;25:498–509.

23. Tossavainen E, Henein MY, Grönlund C, et al. Left Atrial Intrinsic Strain Rate Correcting for Pulmonary Wedge Pressure Is Accurate in Estimating Pulmonary Vascular Resistance in Breathless Patients. Echocardiogr. 2016;33:1156–65.

24. Humbert M, Kovacs G, Hoeper MM, et al. 2022 ESC/ERS Guidelines for the diagnosis and treatment of pulmonary hypertension. The European respiratory journal. 2023;61.

25. Lindholm A, Kjellström B, Seemann F, et al. Atrioventricular plane displacement and regional function to predict outcome in pulmonary arterial hypertension. Int J Cardiovasc Imaging. 2022;38:2235–48.

26. Kurt M, Tanboga IH, Aksakal E, et al. Relation of left ventricular end-diastolic pressure and N-terminal pro-brain natriuretic peptide level with left atrial deformation parameters. European heart journal. Cardiovascular Imaging. 2012;13:524–30.

27. Cameli M, Sparla S, Losito M, et al. Correlation of Left Atrial Strain and Doppler Measurements with Invasive Measurement of Left Ventricular End-Diastolic Pressure in Patients Stratified for Different Values of Ejection Fraction. Echocardiogr. 2016;33:398–405.

28. Bytyçi I, Bajraktari G, Lindqvist P, et al. Compromised left atrial function and increased size predict raised cavity pressure: a systematic review and meta-analysis. Clinical physiology and functional imaging. 2019;39:297–307.

29. Cameli M, Caputo M, Mondillo S, et al. Feasibility and reference values of left atrial longitudinal strain imaging by two-dimensional speckle tracking. Cardiovascular ultrasound. 2009;7:6.

30. Mele D, Pedini I, Alboni P, et al. Anatomic M-mode: a new technique for quantitative assessment of left ventricular size and function. The American journal of cardiology. 1998;81:82G–5G.

31. Gan GCH, Ferkh A, Boyd A, et al. Left atrial function: evaluation by strain analysis. Cardiovascular diagnosis and therapy. 2018;8:29–46.

32. Vollema EM, Amanullah MR, Prihadi EA, et al. Incremental value of left ventricular global longitudinal strain in a newly proposed staging classification based on cardiac damage in patients with severe aortic stenosis. European heart journal. Cardiovascular Imaging. 2020;21:1248–58.

33. Fröjdh F, Soundappan D, Sörensson P, et al. Strain measures of the left ventricle and left atrium are composite measures of left heart geometry and function. medRxiv. 2023;2023.05.04.23289077.

34. Dickinson MG, Lam CS, Rienstra M, et al. Atrial fibrillation modifies the association between pulmonary artery wedge pressure and left ventricular end-diastolic pressure. European journal of heart failure. 2017;19:1483–90.

35. Prada G, Vieillard-Baron A, Martin AK, et al. Echocardiographic Applications of M-Mode Ultrasonography in Anesthesiology and Critical Care. Journal of cardiothoracic and vascular anesthesia. 2019;33:1559–83.

