## Supplemental data for "Mitral annular plane systolic excursion to left atrial volume ratio – a strainless relation with left ventricular filling pressures"

| **Table S1. Principal and/or contributing diagnoses after right heart catheterization** | |
| --- | --- |
| Heart failure | 24 (32.9) |
| Chronic thromboembolic pulmonary hypertension | 8 (11.0) |
| Idiopathic pulmonary arterial hypertension | 7 (9.6) |
| Systemic sclerosis | 7 (9.6) |
| Normal | 7 (9.6) |
| Associated pulmonary arterial hypertension | 7 (9.4) |
| Hypertrophic cardiomyopathy | 3 (4.1) |
| Tricuspid regurgitation | 3 (4.1) |
| Chronic obstructive pulmonary disease | 3 (4.1) |
| Atrial septum defect | 3 (4.1) |
| Hypertension | 3 (4.1) |
| Ischemic heart disease | 2 (2.7) |
| Systemic lupus erythematosus /Mixed connective tissue disease | 2 (2.7) |
| Cardiac amyloid | 2 (2.7) |
| Constrictive pericarditis | 1 (1.9) |
| Sarcoidosis | 1 (1.4) |
| Aortic stenosis | 1 (1.4) |
